# Genetic analysis of functional rare germline variants across 9 cancer types from the DiscovEHR study

**DOI:** 10.1101/2019.12.09.19013334

**Authors:** Manu Shivakumar, Jason E. Miller, Venkata Ramesh Dasari, David Carey, Radhika Gogoi, Dokyoon Kim, on behalf of the DiscovEHR collaboration

## Abstract

Rare variants play an essential role in the etiology of cancer and characterizing rare germline variants that impact the risk of cancer is an ongoing challenge. We performed a genome-wide rare variant analysis using germline whole exome sequencing (WES) data derived from the Geisinger MyCode initiative to discover cancer predisposition variants. The case-control association analysis was conducted by binning pathogenic and likely pathogenic variants in 5,538 cancer patients and 7,286 matched controls in a discovery set and 1,991 cancer patients and 2,504 matched controls in a validation set across nine cancer types. We discovered 87 genes and 106 pathways significantly associated with cancer (*Bonferroni-corrected P* < 0.05) out of which seven genes and 26 pathways replicated from the validation set (*suggestive threshold P* < 0.05). Further, four genes and 21 pathways were discovered to be associated with multiple cancers (*Bonferroni-corrected P* < 0.05). Additionally, we identified 13 genes and two pathways associated with survival outcome across seven cancers (*Bonferroni-corrected P* < 0.05), where two genes, *PCDHB8* and *DCHS2*, were also associated with survival outcome in TCGA data. In summary, we conducted one of the largest pan-cancer association studies using germline data derived from a single hospital system to find novel predisposition genes and pathways associated with nine cancers. Our results can inform future guidelines for germline genetic testing in cancer, which will be helpful in screening for cancer high-risk patients. This work adds to the knowledge base and progress being made in precision medicine.

## Introduction

Cancer is the second most lethal disease in United States with an estimated 1,735,350 new cases and 609,640 deaths in 2018^1^. Cancer is caused by inherited germline variants and acquired somatic mutations. A recent twin study showed ∼33% heritability of cancer across 23 cancer types with a high estimate of 57% for prostate (MIM: 176807), 31% for breast (MIM: 114480), 38% for kidney (MIM: 144700), and 58% for skin melanoma (MIM: 155600)^2^. Germline genetic markers for cancers have been widely studied leading to the discovery of many heritable predisposition genes such as *BRCA1* (MIM: 113705), *BRCA2* (MIM: 600185) and PALB2 (MIM: 610355) in breast cancer, *RB1* (MIM: 614041) in retinoblastoma and *MLH1* (MIM: 120436), *MSH2* (MIM: 609309), *MSH6* (MIM: 600678), and *PMS2* (MIM: 600259) in Lynch syndrome (MIM: 120435). To date, many genome-wide association studies (GWAS) have been conducted and many more variants and genes have been discovered as associated with various cancer types^3-8^. However, a large portion of inherited genetic factors that result in carcinogenesis is still unknown and many studies are being undertaken to discover these genetic variants. For instance, the genetic contribution explained by all variants discovered to date is about 39% in prostate cancer^2; 9^ and 30% in breast cancer^2; 10^.

Since common variants discovered to be associated with multiple cancers have only modest effect size, the missing heritability could be further explained by rare variants. Moreover, rare variants have been known to contribute to various complex diseases including cancer^11-13^. The aggregation of rare variants in a gene can lead to loss of function of the gene or change in expression^14^. Similarly, since pathways perform a sequence of biochemical actions leading to a cellular function or product, changes in the expression of genes involved within a pathway can lead to cancer^15; 16^. Previous studies have also indicated that cancer is caused by an accumulation of a number of singular or rare variants in particular genes or pathways^12^. To that effect, binning the pathogenic and likely pathogenic rare variants into genes and pathways would help us increase statistical power to detect associations and also infer biological mechanisms^13; 17; 18^.

The MyCode community initiative is a precision medicine project, launched at Geisinger in 2007, which enabled the storage of blood, serum, and DNA samples in a system-wide biorepository that is available for use in broad research^19^. To date, over 244,000 patients have signed up for the MyCode initiative and over 90,000 patient blood samples have been sequenced as part of the DiscovEHR project in collaboration with the Regeneron Genetics Center^20^. The sequenced data can be easily linked to the electronic health record (EHR) of the patient, allowing access to rich longitudinal data. Apart from the EHR, Geisinger also maintains a cancer registry that contains all the patients diagnosed or treated for cancer at any Geisinger medical facility. Further, as part of the MyCode program, the genetic data is also being used to detect increased risk of developing one or more of 21 medically actionable conditions, including breast cancer, ovarian cancer (MIM: 167000), Marfan syndrome (MIM: 154700), Lynch syndrome, etc., and the results are returned to the patients though a “Return of Results” program^21^. Moreover, many similar programs around the world are helping to integrate genomics into clinical practice^22^. Manolio et al. emphasize that the integration of genomic findings to clinical practice has been relatively slow and insist on the need to have an openly accessible knowledge base of variants, phenotypes and clinically actionable variants^23^. The sharing of genetic findings is likely to help the scientific research community to improve our understanding of the phenotype of interest and propel precision medicine by bringing more genomics into clinical practice.

In summary, we conducted one of the largest pan-cancer association studies using germline data from a single hospital system to find novel genes and pathways associated with nine cancers. Our study also validates several genes and pathways that have already been implicated in other genome-wide association studies. We identified 87 genes that were significant across cancers, of which seven were replicated in an independent dataset and four genes were shared among multiple cancers. We also identified 106 pathways that reached genome-wide significance, of which 26 pathways were replicated. Further, 21 pathways were significant across multiple cancers. In addition to the genes and pathways associated with cancer risk, we also identified 13 genes and two pathways associated with survival outcome across cancers.

## Results

### Study design and population characteristics

This study was based on a subset of 7,449 cancer cases and 9,792 controls selected by matching age, BMI and gender from ∼90,000 sequenced samples from the DiscovEHR study. The samples were sequenced in two phases using different platforms as described in the Methods section. In phase 1, 60,000 samples were sequenced, and 5,538 cancer patients across nine cancers and 7,286 matched controls were pulled. In phase 2, 30,000 samples were sequenced which included 1,991 cancer patients and 2,504 matched controls. Consequently, the phase 2 dataset was used to replicate results from phase 1. Cancer patient IDs that were retrieved from the cancer registry were classified into particular cancers using International Classification of Diseases for Oncology (ICD-O) codes. After classifying the cancer patients to their respective cancers, only nine cancers, including bladder (MIM: 109800), breast, colorectal (MIM: 114500), kidney, lung (MIM: 211980), melanoma, prostate, thyroid (MIM: 188550), and uterine cancer, had more than 300 samples in the discovery set. A low number of samples in association studies results in a higher type 1 error rate and lower statistical power to detect associations^24^. Thus, the rest of the cancers were excluded from this study. The distribution and basic demographics of patients across these cancers are shown in Table 1. A common set of controls were used for all the cancers except breast, uterine and prostate cancer as they are sex-specific cancers. The sex-matched controls were separately pulled to match the same number of controls across all cancers.

**Table 1.**
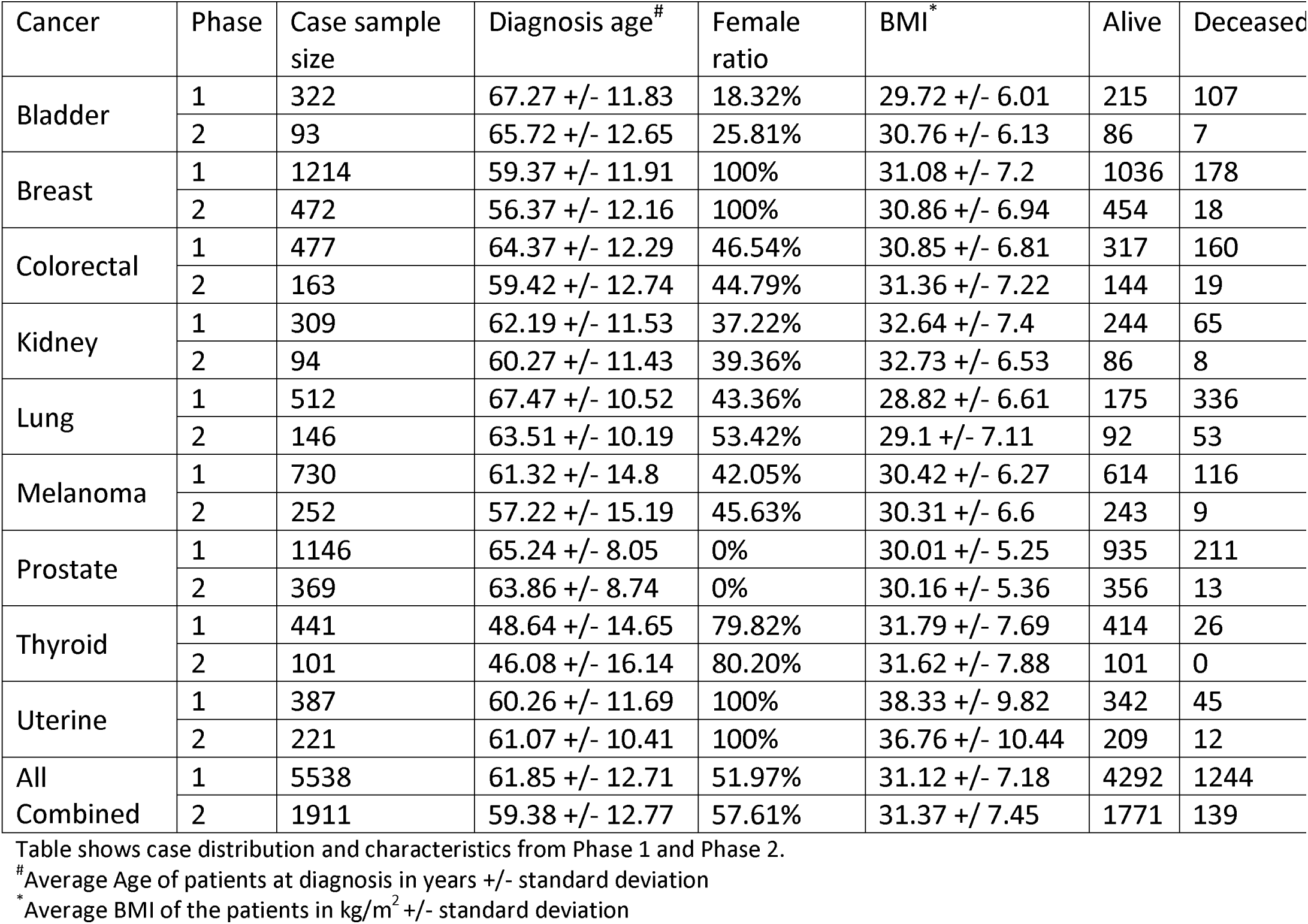
Characteristics for the cancer patients.

Table 1 provides age, BMI, female ratio, and vital status (alive or deceased) information across cancers. Among all cancers, breast cancer had the largest number of cases (N = 1,214) followed by prostate cancer (N = 1,146). Further, uterine cancer had a significantly higher average BMI as compared to other cancers. The average BMI for uterine cancer patients was 38.33 kg/m^2^ in the discovery dataset and 36.76 kg/m^2^ in the replication dataset. Additionally, lung cancer had the highest number of cases who are deceased, which is expected as lung cancer is by far the leading cause of death due to cancer^1^. Further, observing the female ratio across the cancers also shows a gender disparity in some cancers. Specifically, the incidence rate in bladder cancer was found to be 4.46 fold higher in male than female, and in thyroid cancer it was 3.95 fold higher in female than male. The difference in incidence rates have been well documented in other studies with a 3-4 times increased risk of bladder cancer risk in men^25^ and 2.9 times increased risk of thyroid cancer in women^26^.

Variant filtering based on functional annotation and scores improves power and has been successfully used in many association studies^14; 27^. In this study, the variants from whole exome sequence data were annotated using Variant Effect Predictor (VEP)^28^ and ClinVar^29^. Subsequently, only the variants categorized as pathogenic and likely-pathogenic based on the annotations were retained for further analysis. A strategy for classifying variants as pathogenic and likely pathogenic is elaborated in the Methods section. Additionally, all common variants were removed, and only rare variants (MAF < 0.05) were retained. The number of variants available after filtering out for each cancer cohort is listed in Table S1.

### Pathway-based rare variant analysis

Association analysis using rare variants usually suffers from a lack of power as very large datasets are required. Therefore, rare variants are often binned into a biologically informed-unit such as a gene or pathway to improve the power^17^. In this study, pathogenic and likely pathogenic rare variants with minor allele frequency (MAF) < 0.05 were binned into genes followed by KEGG pathways using BioBin^17; 30; 31^. Next, an association test was performed to determine if the gene/pathway is significantly associated with the phonotype. SKAT-O is an optimal unified approach that combines a burden and non-burden sequence kernel association test (SKAT) test, and maintains power regardless of the direction of effect and causality of the variants^32^. After determining the association p-values, they were adjusted for multiple testing in each cancer type separately using a Bonferroni correction. Any genes/pathways with a Bonferroni-corrected *P* < 0.05 were considered as significant results. The same procedure was followed to conduct rare variant analysis in discovery and replication datasets. Figure 2 shows all the pathways that were Bonferroni significant across all cancers and the same are listed in Table S4-S6.

**Figure 1.**
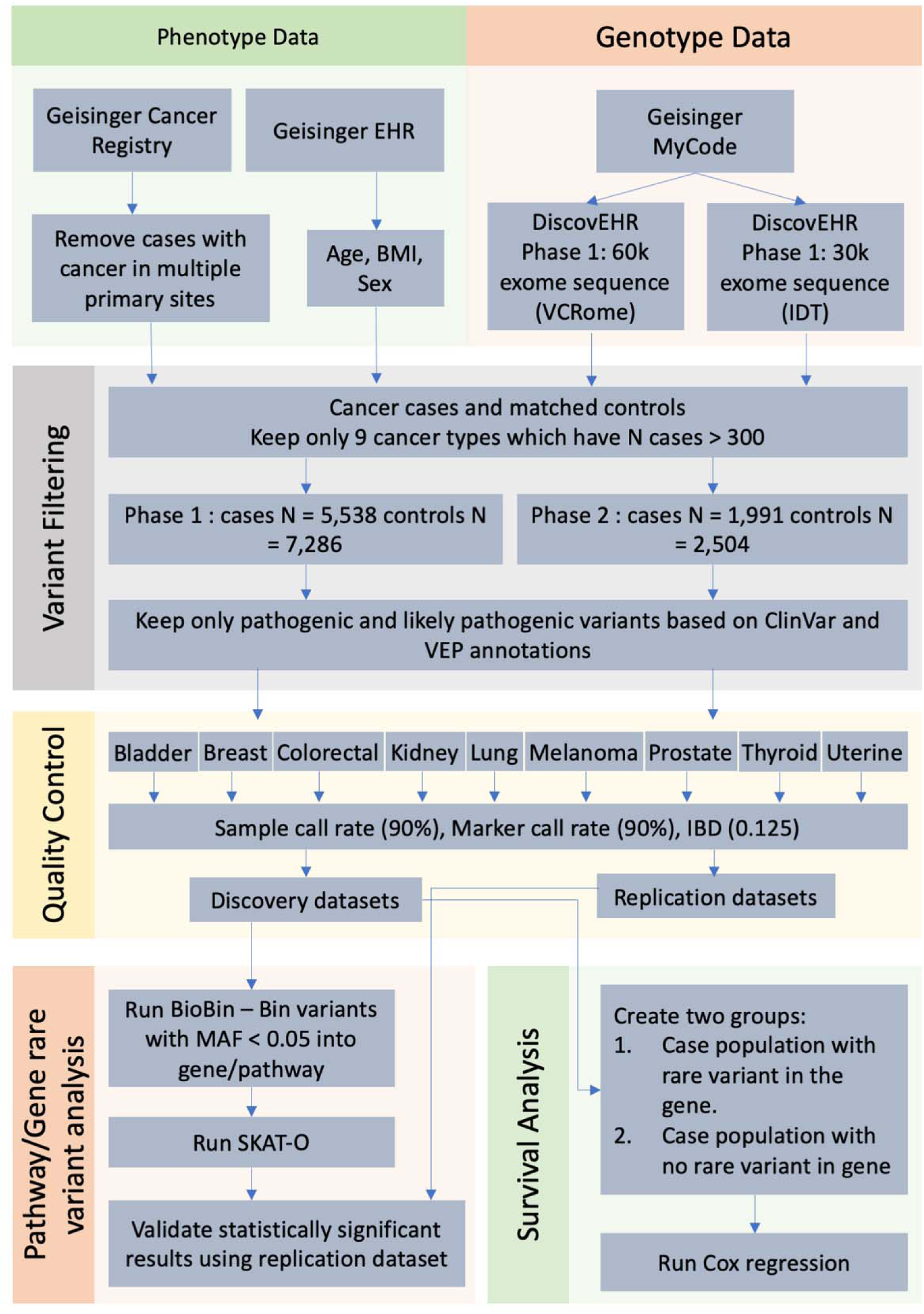
Schematic overview of Pan-caner analysis. The phenotype data was obtained from the Geisinger cancer registry and EHR and the genotype data was obtained from DiscovEHR study. Figure shows multiple steps involved in the analysis – variant filtering, quality control, rare variant analysis, and survival analysis.

**Figure 2.**
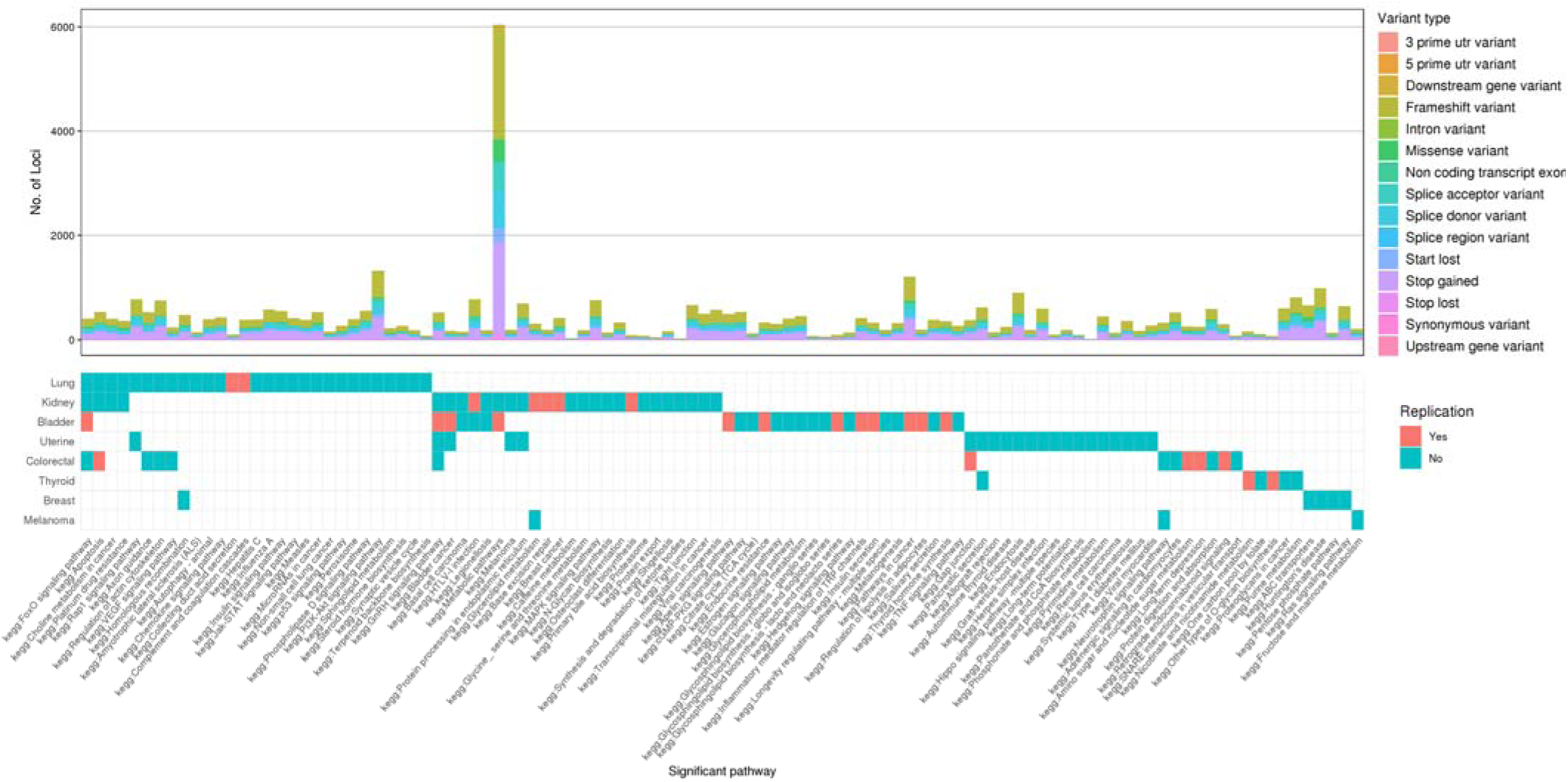
Pathways (x-axis) that were significantly associated with cancer (y-axis) (Bonferroni-corrected *P* < 0.05). The pathways marked as replication ‘Yes’ were significant in discovery (Bonferroni-corrected *P* < 0.05) and replication (SKAT-O *P* < 0.05) datasets. The top bar plot shows the distribution of variant types as annotated by VEP across each pathway.

In total, 106 pathways were found to be significantly associated across all cancers (Figure 2). However, no significant pathways were found in prostate cancer. Further, 26 pathways: 12 in bladder cancer, five in colorectal cancer, five in kidney cancer, two in lung cancer and two in thyroid cancer, marked in red in Figure 2, were replicated from the replication dataset (SKAT-O *P* < 0.05). More information regarding the 26 pathways including locus count, minor allele count (MAC) in cases, MAC in controls, SKAT-O p-value and Bonferroni-corrected p-value in discovery and replication datasets can be found in Table 2. Additionally, 21 pathways were found to be significantly associated in more than one cancer, with the FoxO signaling pathway and GnRH signaling pathway significantly associated with four cancers, followed by apoptosis and bladder cancer significantly associated with three cancers and the rest of the 17 pathways significantly associated with two cancers (Table 3).

**Table 2.** Pathways significantly associated with cancers in both discovery and replication datasets.

| Cancer | KEGG Pathway | Discovery |  |  |  |  | Replication |  |  |  |
| --- | --- | --- | --- | --- | --- | --- | --- | --- | --- | --- |
|  |  | N locus | MAC Case | MAC Control | SKAT-O *P* | Bonferro ni *P* | N locus | MA C Case | MAC Contro l | SKAT- |
| Bladder | Insulin secretion | 325 | 56 | 822 | 2.72E-12 | 8.57E-10 | 160 | 19 | 435 | 2.07E |
| Bladder | Bladder cancer | 124 | 34 | 550 | 4.28E-07 | 1.35E-04 | 55 | 13 | 432 | 1.84E |
| Bladder | GnRH signaling pathway | 358 | 81 | 1728 | 1.99E-06 | 6.28E-04 | 187 | 32 | 761 | 1.47E |
| Bladder | FoxO signaling pathway | 341 | 63 | 1298 | 2.37E-06 | 7.46E-04 | 179 | 23 | 653 | 1.11E |
| Bladder | Inflammatory mediator regulation of TRP channels | 419 | 87 | 1770 | 3.47E-06 | 1.09E-03 | 212 | 31 | 696 | 2.22E |
| Bladder | Pathways in cancer | 1203 | 256 | 5138 | 1.11E-05 | 3.49E-03 | 565 | 100 | 2402 | 2.55E |
| Bladder | Metabolic pathways | 5806 | 1225 | 27356 | 2.23E-05 | 7.04E-03 | 2906 | 425 | 11816 | 3.59E |
| Bladder | Regulation of lipolysis in adipocytes | 191 | 62 | 1215 | 3.45E-05 | 1.09E-02 | 94 | 23 | 427 | 3.08E |
| Bladder | Glycosphingolipid biosynthesis - lacto and neolacto series | 95 | 28 | 601 | 3.88E-05 | 1.22E-02 | 45 | 8 | 196 | 8.40E |
| Bladder | Apelin signaling pathway | 492 | 92 | 1630 | 4.65E-05 | 1.47E-02 | 230 | 39 | 743 | 1.70E |
| Bladder | Endocrine resistance | 325 | 70 | 1435 | 7.95E-05 | 2.50E-02 | 148 | 37 | 984 | 2.92E |
| Bladder | Thyroid hormone synthesis | 352 | 60 | 1100 | 9.94E-05 | 3.13E-02 | 174 | 20 | 473 | 2.05E |
| Colorectal | Gap junction | 269 | 64 | 875 | 1.36E-07 | 4.27E-05 | 128 | 27 | 402 | 8.70E |
| Colorectal | Retrograde endocannabinoid signaling | 296 | 72 | 960 | 1.06E-06 | 3.33E-04 | 150 | 36 | 603 | 7.34E |
| Colorectal | Amino sugar and nucleotide sugar metabolism | 252 | 66 | 947 | 1.06E-05 | 3.35E-03 | 121 | 23 | 321 | 5.19E |
| Colorectal | Long-term depression | 250 | 76 | 999 | 2.25E-05 | 7.10E-03 | 126 | 42 | 570 | 1.27E |
| Colorectal | Apoptosis | 474 | 132 | 1930 | 1.06E-04 | 3.33E-02 | 246 | 85 | 953 | 1.00E |
| Kidney | Base excision repair | 191 | 49 | 865 | 9.83E-10 | 3.10E-07 | 88 | 10 | 242 | 4.25E |
| Kidney | Primary bile acid biosynthesis | 88 | 17 | 199 | 1.60E-05 | 5.03E-03 | 38 | 5 | 70 | 6.18E |
| Kidney | HTLV-I infection | 728 | 248 | 5636 | 2.19E-05 | 6.90E-03 | 342 | 101 | 2260 | 4.55E |
| Kidney | Glycerolipid metabolism | 282 | 74 | 1333 | 7.03E-05 | 2.22E-02 | 152 | 26 | 841 | 1.50E |
| Kidney | Breast cancer | 413 | 60 | 1116 | 8.80E-05 | 2.77E-02 | 184 | 55 | 1137 | 2.62E |
| Lung | Collecting duct acid secretion | 98 | 23 | 166 | 2.29E-06 | 7.22E-04 | 47 | 6 | 104 | 1.11E |
| Lung | Complement and coagulation cascades | 383 | 164 | 2092 | 1.05E-04 | 3.31E-02 | 188 | 22 | 748 | 2.26E |
| Thyroid | Nicotinate and nicotinamide metabolism | 158 | 64 | 942 | 5.73E-05 | 1.80E-02 | 75 | 15 | 389 | 9.20E |
| Thyroid | Other types of O-glycan biosynthesis | 75 | 10 | 134 | 1.22E-04 | 3.85E-02 | 45 | 5 | 57 | 3.10E |

N locus: Total number of genomic loci binned in pathway.
MAC Case: Total minor allele count of variants in pathway in case population.
MAC Control: Total minor allele count of variants in pathway in control population.

**Table 3.** Pathways significantly associated with more than one cancer.

| KEGG Pathway | N Cancers | Cancers |
| --- | --- | --- |
| Apoptosis | 3 | Colorectal, Kidney, Lung |
| Axon guidance | 2 | Colorectal, Lung |
| Basal cell carcinoma | 2 | Bladder, Kidney |
| Bladder cancer | 3 | Bladder, Kidney, Uterine |
| Choline metabolism in cancer | 2 | Kidney, Lung |
| FoxO signaling pathway | 4 | Bladder, Colorectal, Kidney, Lung |
| Gap junction | 2 | Colorectal, Uterine |
| Glycerolipid metabolism | 2 | Kidney, Melanoma |
| GnRH signaling pathway | 4 | Bladder, Colorectal, Kidney, Uterine |
| Homologous recombination | 2 | Breast, Lung |
| HTLV-I infection | 2 | Bladder, Kidney |
| Legionellosis | 2 | Bladder, Kidney |
| Melanoma | 2 | Kidney, Uterine |
| Metabolic pathways | 2 | Bladder, Kidney |
| Neurotrophin signaling pathway | 2 | Colorectal, Melanoma |
| Pancreatic secretion | 2 | Thyroid, Uterine |
| Platinum drug resistance | 2 | Kidney, Lung |
| Protein processing in endoplasmic reticulum | 2 | Kidney, Uterine |
| Rap1 signaling pathway | 2 | Lung, Uterine |
| Regulation of actin cytoskeleton | 2 | Colorectal, Lung |
| VEGF signaling pathway | 2 | Colorectal, Lung |

All pathways were significant in the cancers provided in column 3 with Bonferroni-corrected $P < 0.05$

### Gene-based rare variant analysis

All pathogenic and likely pathogenic variants below MAF < 0.05 were binned into gene boundaries defined by Entrez annotations derived from Library of Knowledge Integration (LOKI) using BioBin^17; 30^. The total number of genes that the variants were binned across all cancers is listed in Table S2. The bar plot in Figure 3 shows the total number of loci binned for a given gene and variant types as annotated by VEP. In total, there were 87 genes that were significantly associated with a specific cancer (Bonferroni-corrected *P* < 0.05) (Figure 3). Furthermore, seven genes - *MLNR* (MIM: 602885), *CPAMD8* (MIM: 608841), *CHRNE* (MIM: 100725), *HOXB13* (MIM: 604607), *SCML4* (HGNC: 256380), *BST1* (MIM: 600387) and *TMEM186* (HGNC: 25880), marked in red in Figure 3, were replicated in the phase 2 dataset (*P* < 0.05) (Table 4).

**Table 4.** Genes associated with cancers that were replicated.

| Cancer | Gene information |  | Discovery |  |  |  |  | Replication |  |  |  |
| --- | --- | --- | --- | --- | --- | --- | --- | --- | --- | --- | --- |
|  | Gene | Chr: Build 38 position | N locus | MAC Case | MAC Control | SKAT-O p-value | Bonferroni corrected p-value | N locus | MAC Case | MAC Control | SKAT-O p-value |
| Kidney | *MLNR* | 13: 49220338-49222377 | 5 | 6 | 39 | 1.07E-07 | 3.70E-04 | 3 | 7 | 48 | 7.89E-04 |
| Kidney | *CPAMD8* | 19: 16892947-17026818 | 23 | 9 | 75 | 2.24E-07 | 7.70E-04 | 14 | 3 | 43 | 1.29E-04 |
| Uterine | *CHRNE* | 17: 4801064-4806369 | 17 | 9 | 95 | 7.92E-07 | 2.70E-03 | 17 | 8 | 27 | 1.29E-04 |
| Prostate | *HOXB13* | 17: 48724763-48728749 | 5 | 19 | 37 | 2.86E-06 | 1.06E-02 | 2 | 8 | 14 | 5.54E-04 |
| Bladder | *SCML4* | 6: 107697297-107845959 | 6 | 5 | 116 | 2.26E-07 | 7.70E-04 | 4 | 3 | 18 | 1.13E-04 |
| Thyroid | *BST1* | 4: 15701866-15774178 | 21 | 5 | 41 | 2.36E-07 | 8.20E-04 | 13 | 4 | 49 | 2.56E-04 |
| Uterine | *TMEM186* | 16: 8795180-8797648 | 3 | 13 | 101 | 8.66E-08 | 3.00E-04 | 2 | 10 | 47 | 4.49E-04 |

N locus: Total number of genomic loci binned in gene. MAC Case: Total minor allele count of variants in gene in case population. MAC Control: Total minor allele count of variants in gene in control population.

**Figure 3.**
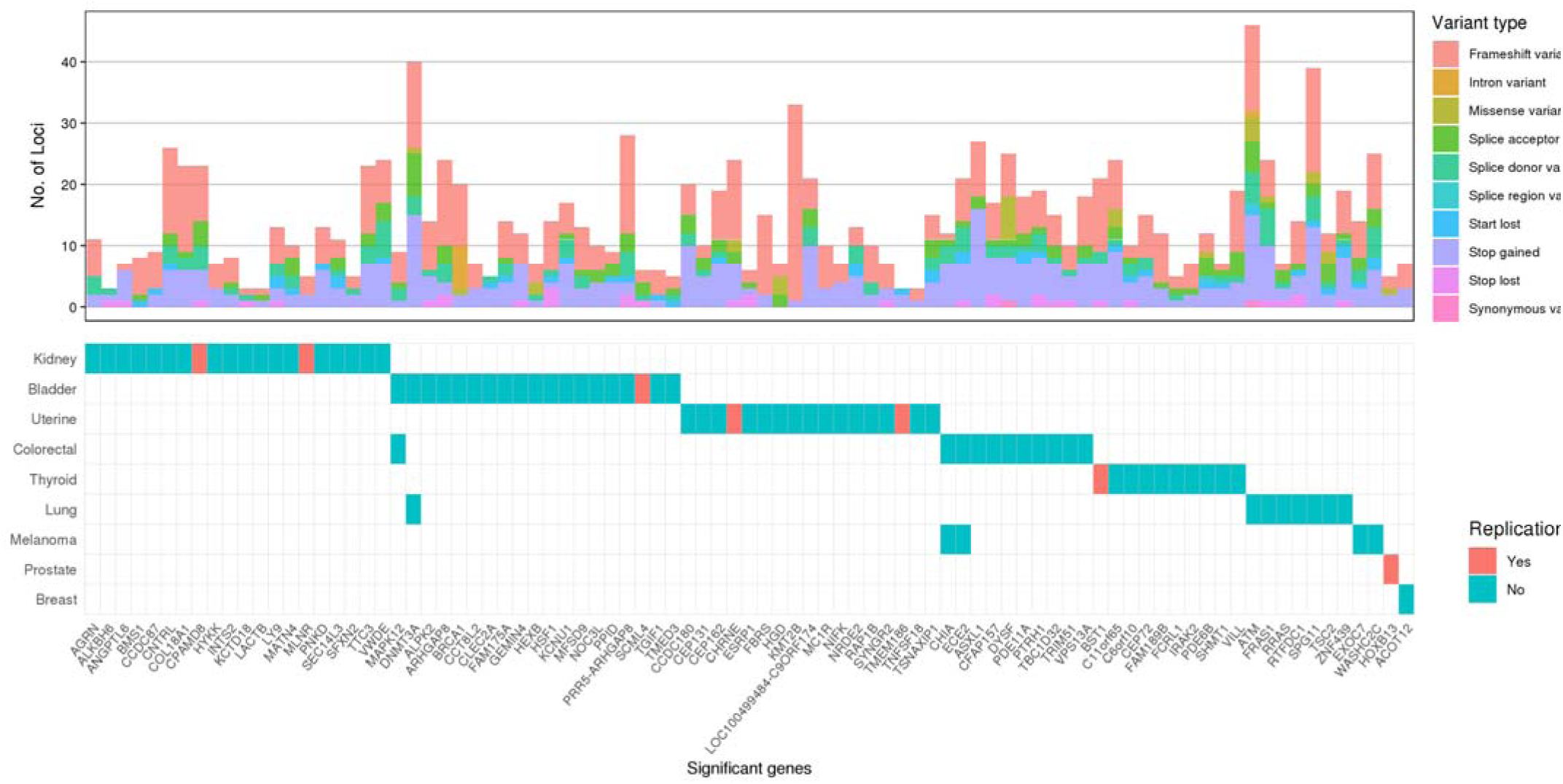
Genes (x-axis) that were significantly associated with cancer (y-axis) using Bonferroni < 0.05. The genes marked as replication ‘Yes’ were significant in discovery (Bonferroni < 0.05) and replication (association p-value < 0.05) datasets. The top bar plot shows the distribution of variant types as annotated by VEP across each gene.

Additionally, four genes, including *MAPK12* (MIM: 602399), *ECE2* (MIM: 610145), *DNMT3A* (MIM: 602769) *and CHIA* (MIM: 606080), were significantly associated in multiple cancers. The gene *MAPK12* was significantly associated with bladder cancer and colorectal cancer, *ECE2* and *CHIA* for melanoma and colorectal cancer, and *DNMT3A* for bladder and lung cancer. Further association test statistics on these genes are described in Table 5. The PhenoGram^33^ plot in Figure 4 shows all the genes found to be significantly associated across all cancers. Additionally, the lollipop^34^ plots in Figure 5 and Figure S1 shows the type of variants – frameshift, missense, stop gained, stop lost, splice acceptor, splice donor, start lost, and their relative position in the gene. Variants that were found in the Catalog of Somatic Mutation in Cancer (COSMIC) database were marked with COSMIC ids. The nonsense stop gained variants marked in yellow usually results in a truncated protein, which are non-functional and the frameshift variants marked in red usually cause a loss of function due to a shift in the reading frame.

**Table 5.** Genes significantly associated with more than one cancer.

| Cancer | Gene information |  | Discovery |  |  |  |  |
| --- | --- | --- | --- | --- | --- | --- | --- |
| Cancer | Gene | Chr: Build 38 position | N locus | MAC Case | MAC Control | SKAT-O p-value | Bonferroni corrected p-value |
| Bladder | *MAPK12* | 22: 50252901-50261810 | 7 | 6 | 42 | 1.18E-06 | 4.02E-03 |
| Colorectal | *MAPK12* | 22: 50252901-50261810 | 9 | 8 | 42 | 1.18E-05 | 4.08E-02 |
| Colorectal | *ECE2* | 3: 184276011-184293031 | 17 | 4 | 17 | 7.23E-06 | 2.50E-02 |
| Melanoma | *ECE2* | 3: 184276011-184293031 | 20 | 6 | 17 | 2.93E-06 | 1.04E-02 |
| Lung | *DNMT3A* | 2: 25232961-25342590 | 30 | 10 | 28 | 6.62E-10 | 2.00E-06 |
| Bladder | *DNMT3A* | 2: 25232961-25342590 | 25 | 6 | 27 | 2.21E-06 | 7.53E-03 |
| Colorectal | *CHIA* | 1: 111290852-111320566 | 9 | 9 | 70 | 4.14E-10 | 1.00E-06 |
| Melanoma | *CHIA* | 1: 111290852-111320566 | 9 | 8 | 70 | 1.38E-05 | 4.89E-02 |

**Figure 4.**
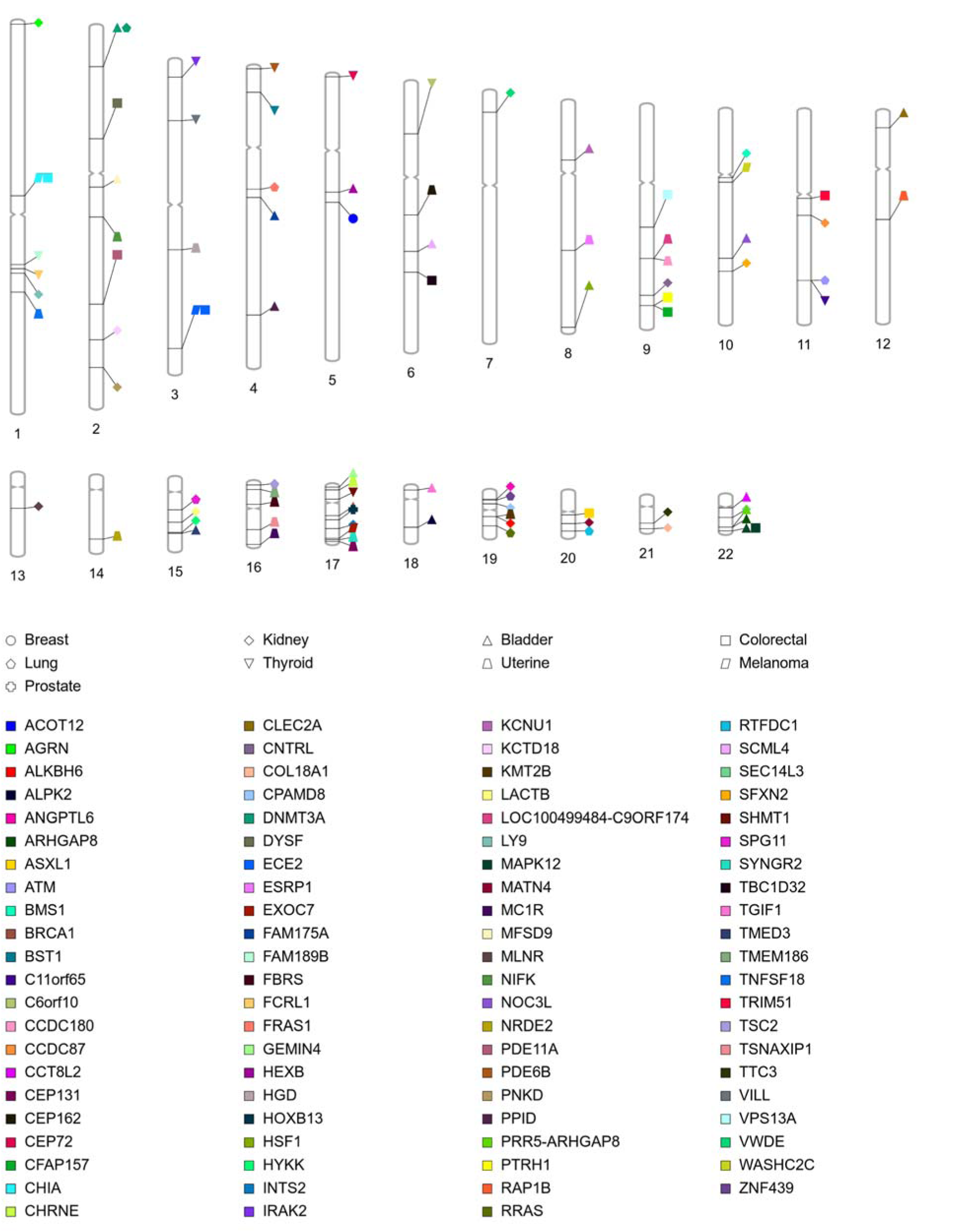
PhenoGram plot of all significant genes across all cancers.

**Figure 5.**
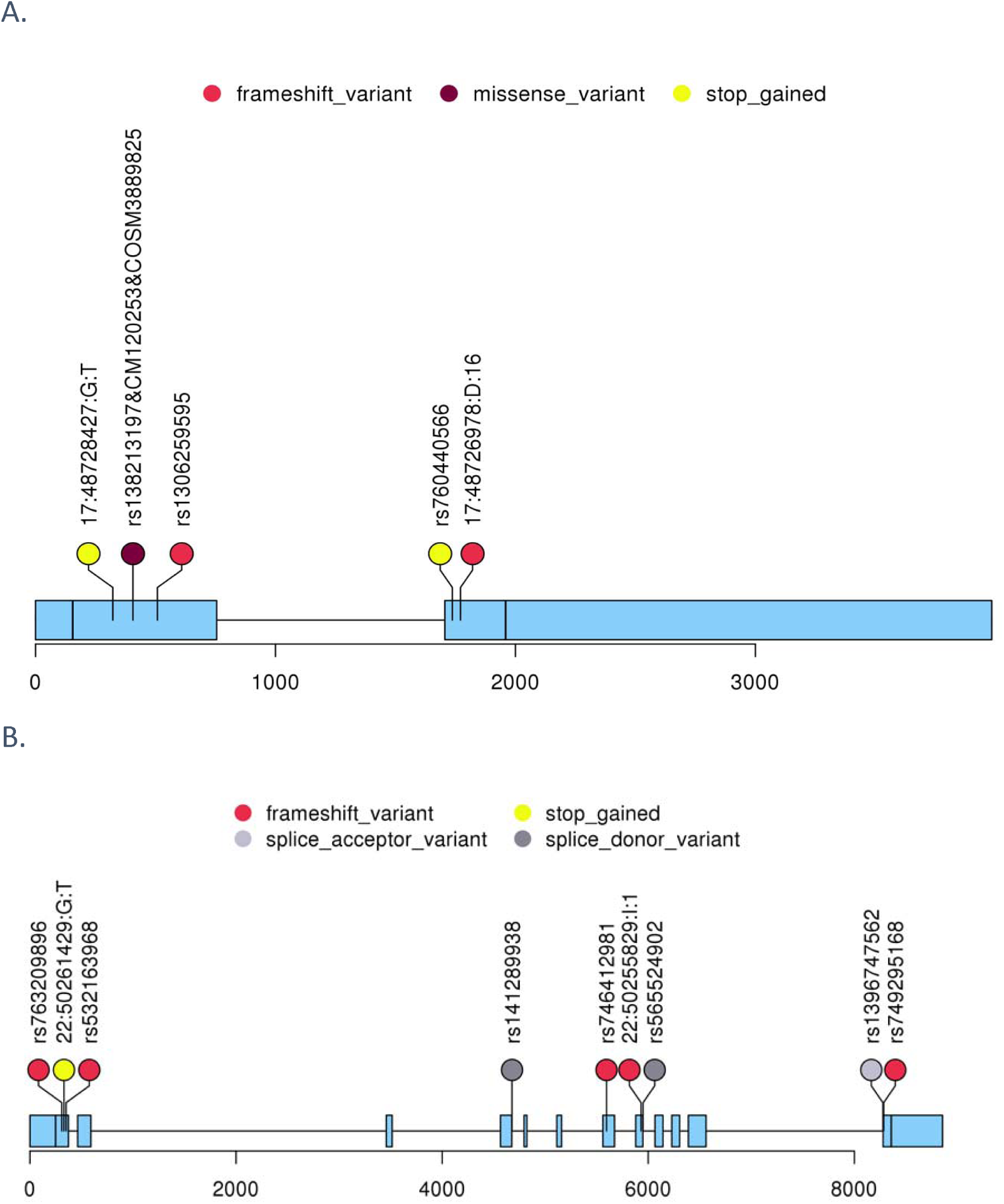
Lollipop plot of genes with all loci binned in them. The color represents different types of variants as assonated by VEP. A. Lollipop plot for HOXB13 gene; B. Lollipop plot for MAPK12 gene; C. Lollipop plot for ECE2 gene.

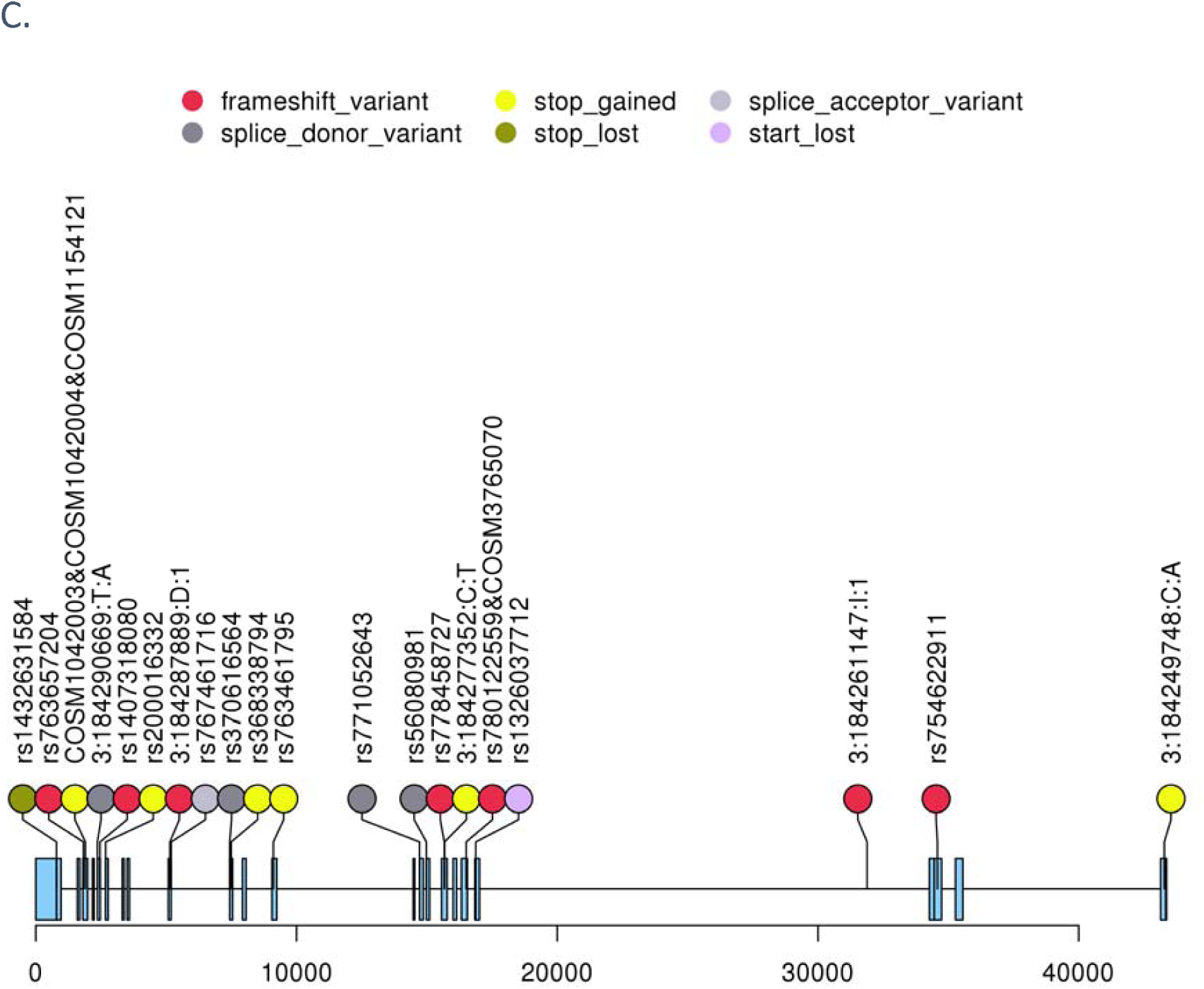

### Pathogenic and likely pathogenic variants in known oncogenes and tumor suppressor genes

Previously, Huang et al.^14^ identified potential genes in cancers with a higher enrichment of pathogenic or likely pathogenic variants identified in the Exome Aggregation Consortium (ExAC) non-TCGA cohort from a curated list of genes that contribute to cancer susceptibility. They identified 28 cancer gene associations (FDR < 0.05) and 16 suggestive associations (FDR < 0.15) by conducting total frequency test (TFT)^14^ on germline data across 33 cancers. We wanted to see if any of these genes also had higher mutational germline burden in our study as it would help validate cancer susceptibility genes and even discover new gene associations that were not statistically significant in Huang et al. As such, the list of gene-cancer pairs were filtered to the nine cancer types under consideration in this study and TFT was run on the genes to validate the enrichment of pathogenic and likely pathogenic variants in cancer patients against control group. Seven genes – *ATM* (lung, MIM: 607585), *CHECK2* (breast, HGNC: 11200), *MSH2* (colorectal), *BRCA2* (thyroid), *POLE* (kidney, MIM: 174762), *PALB2* (breast), *MLH1* (colorectal) were found to be significant at a TFT p-value < 0.05. Moreover, two of the genes – *ATM* (lung) and *BRCA2* (thyroid) found to be previously significant ^14^ were replicated in this study. Further, three genes – *ATM* (lung), *CHECK2* (breast) and *MSH2* (colorectal) were significant at FDR < 0.15. The carrier frequency of these genes and distribution of pathogenic and likely pathogenic variants across oncogenes and tumor suppressor genes is shown in Figure 6 (Table S8). In summary, we were able to validate two genes (TFT p-value < 0.05) and we discovered two more genes with a suggestive association (FDR < 0.15).

**Figure 6.**
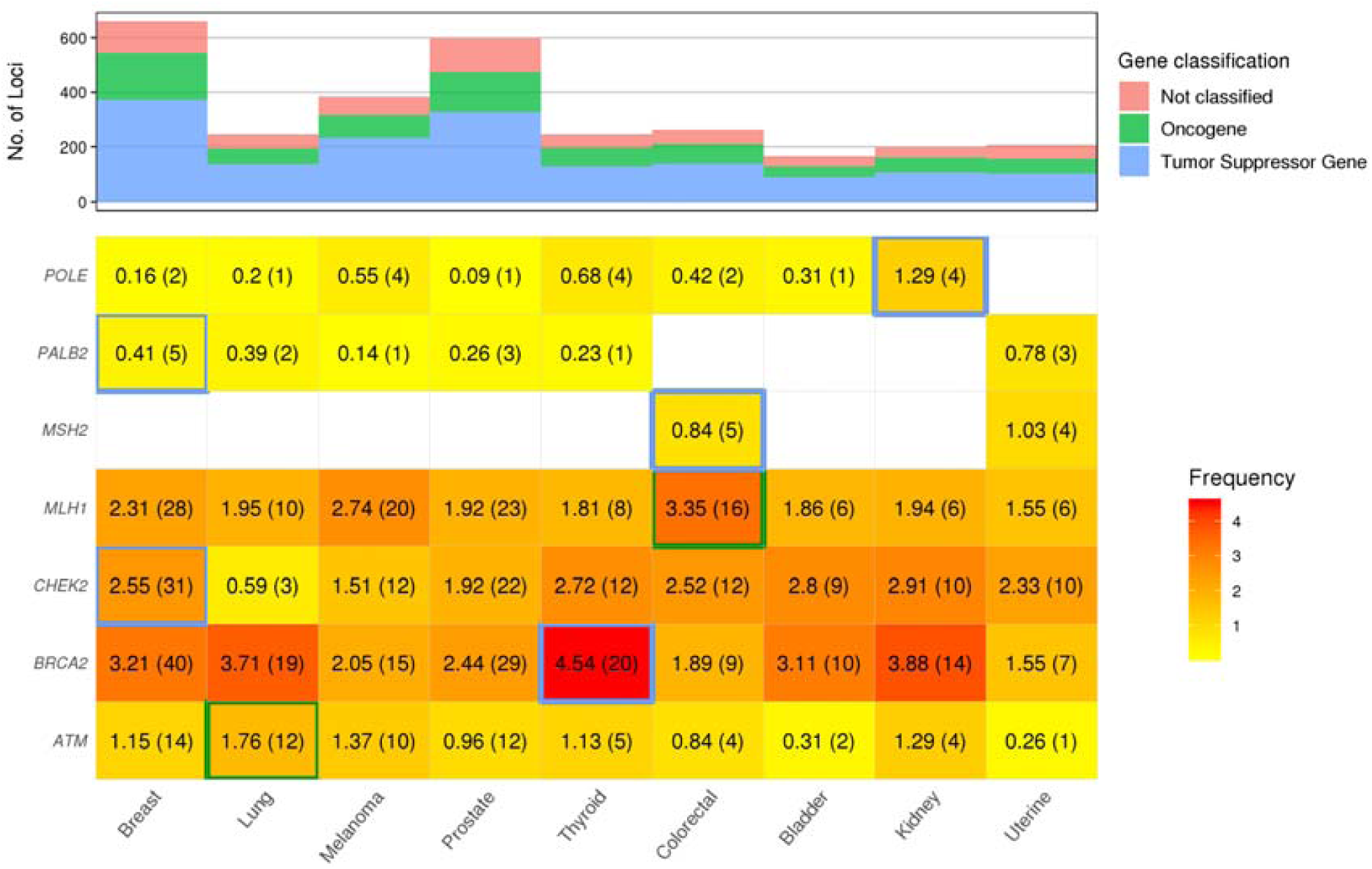
Carrier frequency of pathogenic and likely pathogenic variants across known oncogenes and tumor suppressors. The color of the boxes represents the carrier frequency of pathogenic and likely pathogenic variants as denoted by the frequency legend. The 7 genes significant at TFT p-value < 0.05 are marked with blue or green border. The 2 genes replicated from Huang et al ^14^ are marked with green border.

**Figure 7.**
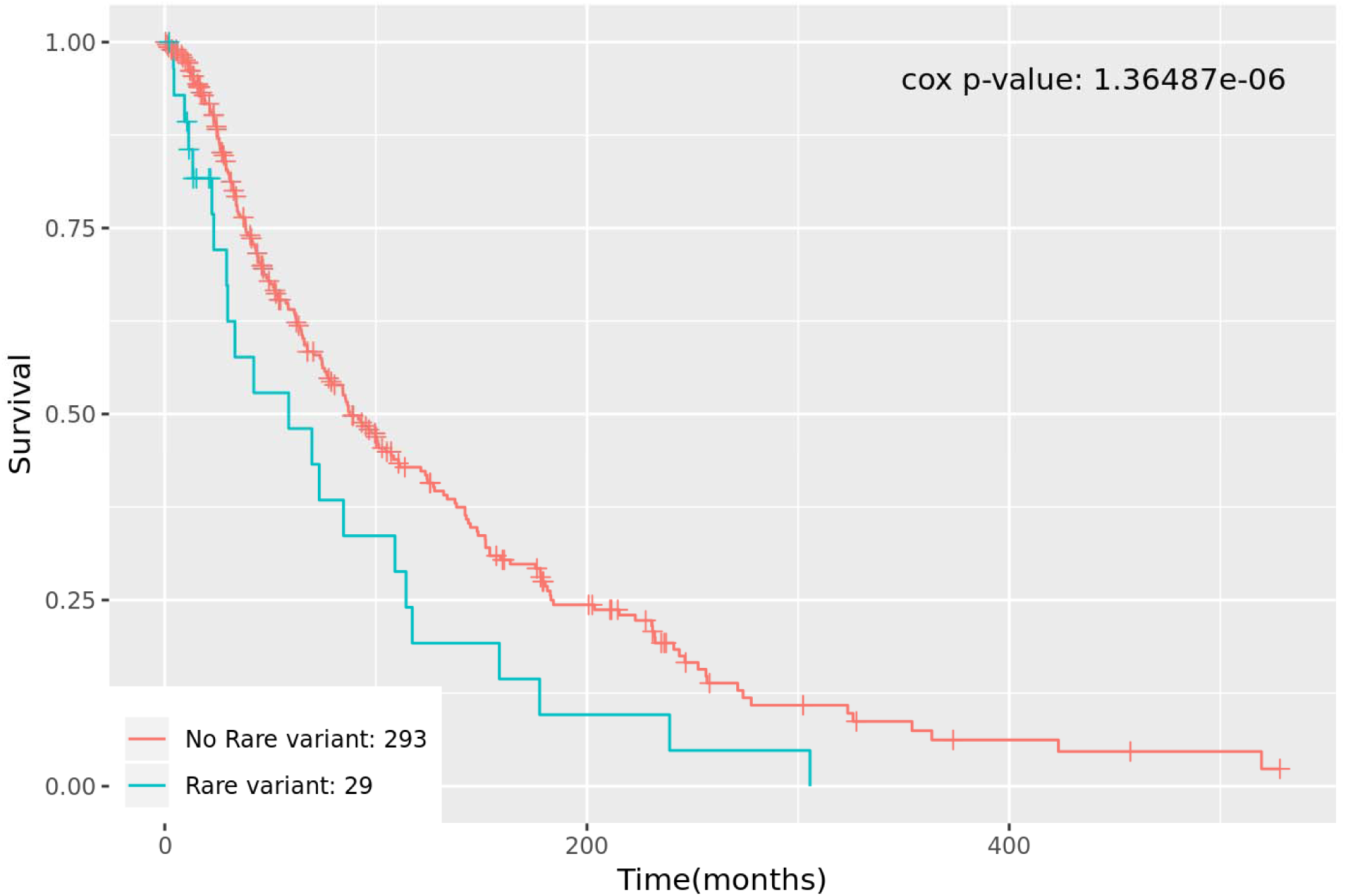
Kaplan Meier survival plot for gene *PCDHB8* which was significantly associated with survival in bladder cancer. *PCDHB8* was also significantly associated with survival in bladder cancer in TCGA dataset.

### Survival analysis

In this study, we also sought to discover variants that have an impact on the survival of patients. To this aim, a weighed burden matrix was used to run cox regression adjusting for age and BMI as covariates. The cox p-values were further adjusted using a Bonferroni correction separately on each cancer type to account for multiple testing. Thirteen genes and two pathways across seven cancers were found to be significantly associated with survival at Bonferroni < 0.05. Further, to confirm that the p-values were not a random effect, permutation testing was conducted by randomly shuffling the weighed burden values among patients and running cox regression 100,000 times. We observed considerably less significant permutation p-values across all genes and pathways. The permutation p-values and other statistics are listed in Table 6 for significant genes and Table 7 for significant pathways. All the Kaplan-Meier survival curves are shown in Figure S2. The genes that were significantly associated were further tested for association with survival in The Cancer Genome Atlas (TCGA) provisional data on the cBio Cancer Genomics Portal (http://cbioportal.org)^35^.The two groups were formed using somatic mutations and mRNA expression (RNA Seq V2 RSEM) using a z-score threshold ± 2. Two genes *PCDHB8* (*Logrank P* = 9.22E-03, MIM: 606334) and *DCHS2* (*Logrank P* = 0.036, MIM: 612486) were significant at a *Logrank P* < 0.05.

**Table 6.**
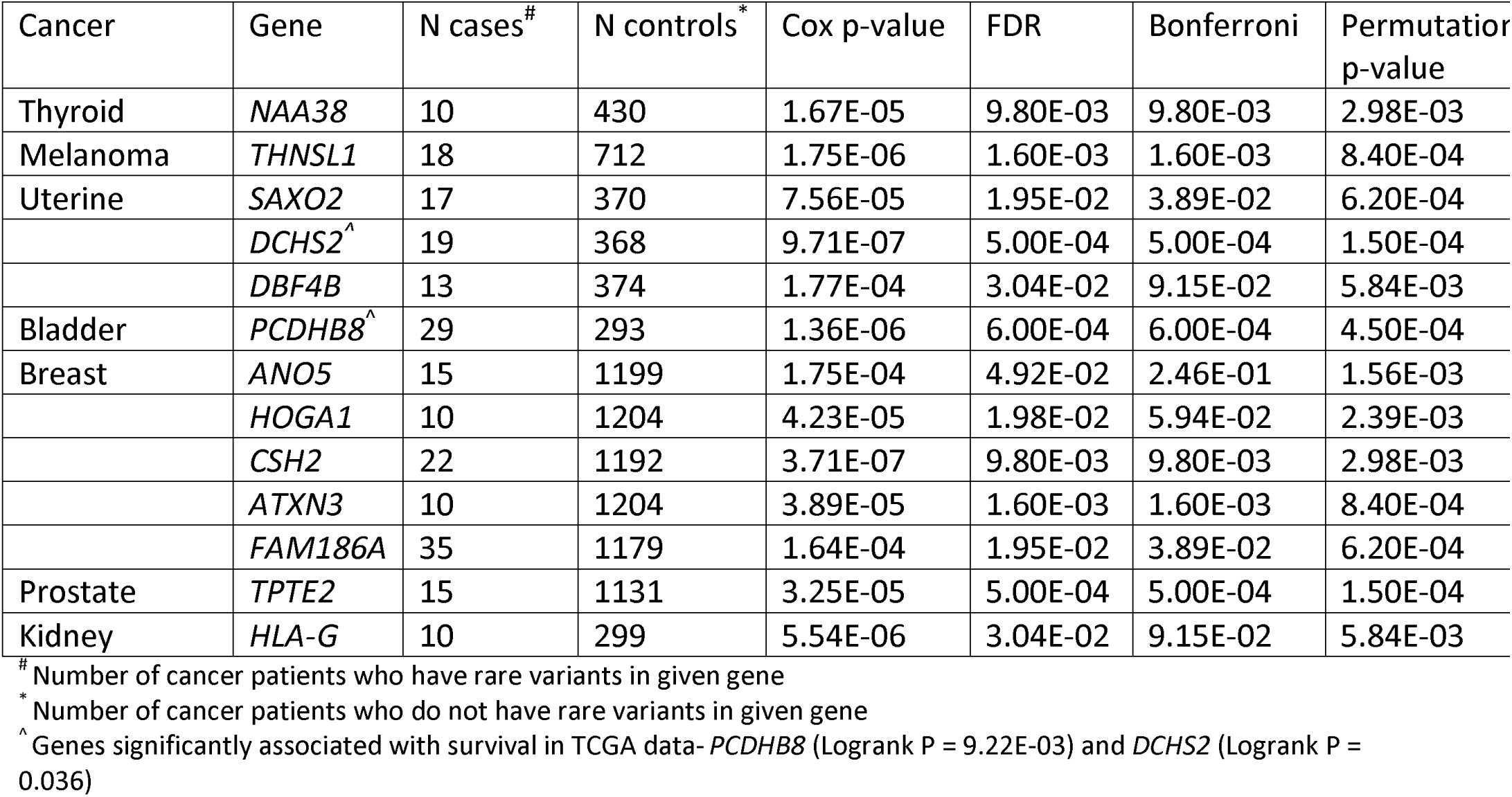
Genes associated with survival.

**Table 7.** Pathways associated with survival.

| Cancer | KEGG Pathway | N cases<sup>#</sup> | N controls<sup>*</sup> | Cox p-value | FDR | Bonferroni | Permutation p-value |
| --- | --- | --- | --- | --- | --- | --- | --- |
| Melanoma | Phenylalanine tyrosine and tryptophan biosynthesis | 19 | 711 | 6.77E-05 | 2.05E-02 | 2.05E-02 | 5.20E-04 |
| Melanoma | Phenylalanine metabolism | 31 | 699 | 1.57E-04 | 2.39E-02 | 4.77E-02 | 3.90E-04 |

<sup>#</sup> Number of cancer patients who have rare variants in given gene
<sup>\*</sup> Number of cancer patients who do not have rare variants in given gene

## Discussion

In this study, we present results from a rare variant analysis conducted across nine cancers using a cohort of 7,449 cancer cases and 9,792 controls from a single hospital system using whole exome sequencing data and clinical data from a patient EHR. A total of 133 pathways (26 replicated) and 91 genes (7 replicated) were identified as associated with cancers. Furthermore, 21 pathways and four genes were associated with multiple cancer types. Additionally, we identified 13 genes and two pathways as associated with survival across multiple cancers.

Many KEGG pathways identified in this study have already been implicated in cancer, such as “pathways in cancer”, “GnRH signaling pathway” ^36^, “bladder cancer”, “FoxO signaling pathway” ^37^, “metabolic pathways”, “gap junction” ^38^, “apoptosis”, “base excision repair”, “melanoma”, “choline metabolism in cancer” and “basal cell carcinoma”. One pathway of interest that is associated with bladder cancer is the “insulin secretion pathway”. Previous studies have shown diabetes mellitus increases the risk of bladder cancer^39^ and is possibly due to administration of the anti-diabetic drug Pioglitazone^40^. Another pathway “HTLV-I infection” was found to be associated with kidney cancer and was also replicated. HTLV-I is a known oncovirus that causes cancer^41^. Further studies on the “HTLV-I infection” pathway could elucidate the role of germline variants in cancers. Another pathway “Legionellosis” was found to be associated with kidney and bladder cancer, Legionella pneumonia in cancer has a very high mortality rate ∼31%^42^, and variants in pathway could play a role in susceptibility or recovery of the patients. Another pathway, the “Hippo signaling pathway” was found to be associated with uterine cancer. The “Hippo tumor suppressor pathway” is known to phosphorylate YAP and TAZ which are critical for cell growth, reprogramming and development^43^. The Hippo pathway also interacts with the PI3K/AKT pathway which is commonly involved in cancer^43^. Additionally, Hippo pathway is known to affect the survival of cancer patients^44^ and in this study, one of the genes *DCHS2*, which is part of Hippo pathway, was also associated with survival in uterine cancer and it was also replicated in the TCGA data.

A number of previous studies have shown *HOXB13* to be associated with prostate cancer^45-48^, and in this study as well, *HOXB13* was found to be associated with prostate cancer in the discovery dataset and was replicated. Another gene, *CPAMD8*, which is involved in broad-spectrum protease inhibition, innate immunity and damage control was found to be associated with kidney cancer in the discovery and replication datasets^49^. *CPAMD8* is known to be substantially expressed in kidney^49; 50^ and given its functional role, rare gene-disruptive variants in *CPAMD8* could lead to carcinogenesis. We also identified two genes associated with uterine cancer that replicated - *CHRNE* which is a subunit of nicotinic acetylcholine receptors (nAChRs) and *TMEM186* which is a member of the transmembrane protein family. Nicotine, a compound present in cigarettes, mediates cell proliferation and angiogenesis though nicotinic acetylcholine receptors (nAChRs) and its subunits^51^. Still, its mechanism of action is not well understood for uterine cancer where some studies have shown smoking to reduce the risk of uterine cancer contrary to other cancers^51; 52^. Again, the exact role of *TMEM186* in uterine cancer is also unexplored. TMEMs are differentially regulated in many types of cancers and some TMEMs are known to act as tumor suppressors while others as oncogenes^53^. Further, in bladder cancer, the Putative Polycomb group (PcG) protein gene (*SCML4*), is involved in the regulation of crucial developmental and physiological processes and is known to promote proliferation and inhibit apoptosis. *SCML4* was replicated in bladder cancer^54^.

Different cancer types share some pathways and genes, which generally include common tumor suppressor genes and oncogenes^14^. In this study, we identified 21 pathways and four genes that were associated with multiple cancers. Two of the genes *MAPK12* and *DNMT3A* are well known genes involved in cancer with *MAPK12* acting as *p38 MAPK*, which is involved in cell differentiation, apoptosis and autophagy, whereas *DNMT3A* is involved in DNA methylation and its disruption leads to tumorigenesis^55; 56^. Gene *ECE2* cleaves endothelin-1 (ET-1) which is a potent vasoconstrictor peptide and ET-1 is known to be involved in angiogenesis, apoptosis and growth in colorectal cancer and melanoma^57^. Elevated levels of plasma levels and increased immunopositivity of ET-1 has been observed in colorectal cancer^57^.

We also discovered 13 genes that are associated with survival in cancers. The presence of rare pathogenic and likely pathogenic variants reduced the overall survival rate for all the genes identified across cancers. Some of the genes discovered were already known to be associated with survival in cancers and a subset of them have also been suggested as a target for cancer therapy like *SAXO2* (HGNC: 283726) which is involved in microtubule binding in uterine cancer^58^, *PCDHB8* whose downregulation is known to result in poor prognosis in bladder cancer^59^, *ATXN3* (MIM: 607047) whose downregulation increases expression of tumor suppressor *PTEN* (MIM: 601728)^60^, and *TPTE2* (MIM: 606791), a homolog of *PTEN*, whose upregulation suppresses metastasis and/or tumorigenesis^61^. Other genes were associated with survival in this study - *ANO5* (MIM: 608662) is known to regulate cell migration and invasion^62^, the *HOGA1* (MIM: 613597) gene is involved in metabolism, *CSH2* (MIM: 118820) is involved in postnatal and intrauterine growth^63^ and *HLA-G* (MIM: 142871) offers an immune escape mechanism as it is involved in cytokine signaling in the immune system and class I MHC mediated antigen processing and presentation^64^. Thus, disruption of the normal activity of these genes could promote cancer. Another gene *NAA38* (MIM: 617990)which was associated with survival in thyroid has been shown to be associated with survival in glioblastoma (MIM: 137800)^65^. Furthermore, two genes *PCDHB8* and *DCHS2* were significantly associated with survival in the TCGA provisional dataset.

Even though many associations were identified in this study, further studies would be required to elucidate the molecular mechanisms. A shortcoming of this study was that the replication cohort was underpowered to replicate all the findings. Additionally, the participants in the replication cohort were derived from participants who enrolled into the MyCode program more recently than the discovery dataset. Therefore, most of the patients in the replication set were alive and it was not practical to run survival analysis using the replication dataset. The limitations imposed by the sample size and power would be addressed in the future as the MyCode and DiscovEHR programs are still ongoing and more samples are being sequenced. Another limitation of our study is that our population predominantly consists of European ancestry, mainly due to the patient population at Geisinger which is predominantly of European ancestry.

In conclusion, this study conducted genome-wide rare-variant analysis to find novel genes and pathways associated across nine cancers. We also replicated many genes and pathways that are very well known in cancers, which further emphasizes the fact that some portion of the missing heritability is attributed to rare variants. We also identified some genes associated with the survival of the patients which have already been suggested as targets in cancer therapy. We also discovered novel genes and pathways associated with survival of patients which could be potential targets for cancer therapy. The genes and pathways discovered in this study can be used to screen for high-risk cancer patients and personalized therapy. In summary, results from this study could help define a portion of the missing heritability associated with cancer and have broad applications in precision medicine.

## Methods

### Study population

The study population consisted of Geisinger patients who consented to participate in the MyCode community initiative. As part of MyCode initiative, individuals agreed to provide blood and DNA samples for broad, future research, including genomic analyses as part of the Regeneron-Geisinger DiscovEHR collaboration and linking to data in the Geisinger EHR under a protocol approved by the Geisinger Institutional Review Board. The cases were a subset of cancer patients from a cancer registry that were part of 90,000 patients sequenced as part of the DiscovEHR project. The cases were classified into different cancers using ICD-O site codes as defined in Table S3. Cases that were recorded as having cancer in multiple primary sites were removed. Further, only cancers with at least 300 cases in 60,000 patients were sequenced in phase 1 of the DiscovEHR study were included and other cancers were discarded due to a low number of cases. A common control set was selected for all non-sex specific cancers using matched age and BMI to cases from a pool of patients who did not have any ICD9/ICD10 code related to cancer in a problem-list entry of the diagnosis code, an inpatient hospitalization-discharge diagnosis code, or an encounter diagnosis code. The controls for breast, uterine and prostate cancer were pulled separately to have the same number of controls as the common control set. Age was calculated as age at diagnosis for cases and current age if alive or age at death for controls. The median of BMI values recorded in the EHR from a year before diagnosis date was used as BMI for cases. Furthermore, the median of BMI values from a year before current date or date of death for alive and deceased patients respectively was used as BMI for controls.

### Sequencing and quality control

All the study population was sequenced as part of the DiscovEHR project at the Regeneron Genetic Center. Initially, around 60,000 samples were sequenced using NimbleGen probe target-capture (SeqCap VCRome) and further a separate batch of 30,000 samples were sequenced using xGen capture (Integrated DNA Technologies) followed by sequencing on the Illumina HiSeq 2500. The variant calling was done using GATK^66; 67^. Further detailed description of sequencing is available at Shivakumar et al.^13^ and Mirshahi et al.^68^. Additional call rate quality controls were applied. The markers and samples with a call rate below 90% were filtered out. All related patients showing up to 3^rd^ degree relatedness corresponding to IBD > 0.125 were removed.

#### Variant annotation and filtering

All variants were annotated using VEP and ClinVar. All the loci that satisfied the following two conditions were retained and the rest were filtered out.

1. Loci that were annotated with impact ‘HIGH’ using VEP.
2. Loci that were annotated as pathogenic and likely pathogenic with at least 1 star using ClinVar.

Variants that satisfied the conditions were considered pathogenic and likely pathogenic and all analysis were run using only these loci.

#### Gene based rare variant association

All the variants that were annotated as pathogenic and likely pathogenic were binned using BioBin^17; 30^. BioBin uses pre-compiled knowledge in a LOKI database, which is compiled using information from various data sources including Entrez and KEGG. The variants were binned into genes using Entrez annotations. Only variants with MAF < 0.05 were considered rare and the rest of the variants were filtered out. Additionally, bins with less than 20 variants (MAC) were filtered out due to low sample size. Further, the binned variants were weighed using Madsen-browning weights^69^. The statistical association tests were run using SKAT-O implemented as R package^32^. Additionally, the association tests were adjusted using age, BMI and first four principle components as covariates. The principle components were calculated using EIGENSOFT^70^, using common variants after LD pruning with indep-pairwise 50 5 0.5 and Hardy-Weinberg equilibrium of 10^−6^. The association test p-values were further adjusted using Bonferroni correction to account for multiple testing correction.

#### Pathway based rare variant association

BioBin was used to bin rare variants with a MAF < 0.05 into KEGG pathways derived from LOKI^17; 30^. Any pathway bin that did not contain a total of at least 20 variants across case and cancer were filtered out due to small sample size. Further all the bins were weighted using Madsen-browning weighting^69^. Statistical association was run using SKAT-O implemented as R package ^32^. The association tests were adjusted using age, BMI and four principle components as covariates. Further, the association p-values were adjusted using Bonferroni correction.

#### Survival analysis

Survival analysis was run using cox regression adjusting for age and BMI. Specifically, the BioBin bin-phe output files which contains a weighted burden of variants were used to get the weighted burden of each patient for the bin (gene/pathway) and cox regression was run on the bin adjusting for age and BMI. Survival analysis was performed on each cancer using gene-based bins and pathway-based bins. Further, survival p-values were adjusted for multiple testing using Bonferroni correction.

## Data Availability

The summary data supporting the conclusions of this manuscript will be made available by the authors to any qualified researcher subject to a data use agreement.

## Author contributions

MS, JEM, VRD, RG and DK designed and conceived the project. MS and JEM carried out the methodology and implementation. DK and RG helped supervise the project. MS wrote the paper in consultation with JEM, VRD, RG and DK.

## Acknowledgement

This work was supported by NLM R01 NL012535. This project was also funded, in part, under a grant with the Pennsylvania Department of Health (#SAP 4100070267). The Department specifically disclaims responsibility for any analyses, interpretations or conclusions. We gratefully acknowledge the funding support from Geisinger Medical Center (SRC-075) (RG) and Rice Women’s Cancer Research Fund (RG and VRD). Support for this work was also provided by NHGRI T32HG009495-01 (JEM). The funders specifically disclaim responsibility for the study design, data collection, analyses, interpretation, conclusions, and writing of the manuscript.

## Web resources

BioBin, https://ritchielab.org/software/biobin-download

VEP, https://ensembl.org/info/docs/tools/vep/index.html

ClinVar, https://www.ncbi.nlm.nih.gov/clinvar/

EIGENSOFT, https://www.hsph.harvard.edu/alkes-price/software/

OMIM, http://www.omim.org/

R statistical software, https://www.r-project.org/

NCBI, https://www.ncbi.nlm.nih.gov/gene/4049

DiscovEHR, http://www.discovehrshare.com/

